# Estimating the case fatality ratio of the COVID-19 epidemic in China

**DOI:** 10.1101/2020.02.17.20023630

**Authors:** Xing Wang, Zihui Ma, Yi Ning, Chen Chen, Rujin Chen, Qiwen Chen, Heng Zhang, Chunming Li, Yan He, Tao Wang, Cheng Tong, Junqing Wu, Yuyan Li, Handong Ma, Shaodian Zhang, Hongxin Zhao

## Abstract

**Background:** Corona Virus Disease 2019 (COVID-19) due to severe acute respiratory syndrome coronavirus 2 (SARS-CoV-2) emerged in Wuhan city and rapidly spread throughout China since late December 2019. Crude case fatality ratio (CFR) with dividing the number of known deaths by the number of confirmed cases does not represent the true CFR and might be off by orders of magnitude. We aim to provide a precise estimate of the CFR of COVID-19 using statistical models at the early stage of the epidemic.

**Methods:** We extracted data from the daily released epidemic report published by the National Health Commission P. R. China from 20 Jan 2020, to 1 March 2020. Competing risk model was used to obtain the cumulative hazards for death, cure, and cure-death hazard ratio. Then the CFR was estimated based on the slope of the last piece in joinpoint regression model, which reflected the most recent trend of the epidemic.

**Results:** As of 1 March 2020, totally 80,369 cases were diagnosed as COVID-19 in China. The CFR of COVID-19 were estimated to be 70.9% (95% CI: 66.8%-75.6%) during Jan 20-Feb 2, 20.2% (18.6%-22.1%) during Feb 3-14, 6.9% (6.4%-7.4%) during Feb 15-23, 1.5% (1.4%-1.6%) during Feb 24-March 1 in Hubei province, and 20.3% (17.0%-25.3%) during Jan 20-28, 1.9% (1.8%-2.1%) during Jan 29-Feb 12, 0.9% (0.8%-1.1%) during Feb 13-18, 0.4% (0.4%-0.5%) during Feb 19-March 1 in other areas of China, respectively.

**Conclusions:** Based on analyses of public data, we found that the CFR in Hubei was much higher than that of other regions in China, over 3 times in all estimation. The CFR would follow a downwards trend based on our estimation from recently released data. Nevertheless, at early stage of outbreak, CFR estimates should be viewed cautiously because of limited data source on true onset and recovery time.

## Introduction

On December 8, 2019, the first pneumonia cases with unknown etiology was identified in Wuhan city, Hubei province, China [1]. Subsequently, World Health Organization (WHO) appointed an expert group arrived in China to guide the epidemic management. On January 8, 2020, the pathogen of this unexplained infected pneumonia was isolated by Chinese researchers, now known as severe acute respiratory syndrome coronavirus 2 (SARS-CoV-2) [2-4], which resembled to the severe acute respiratory syndrome coronavirus (SARS-CoV) [1]. On January 25, 2020, WHO released the Novel Coronavirus (2019-nCoV) situation report-5 and stated that the new coronavirus epidemic has posed a very high risk to China and a high risk to both regional level and global level [5]. On February 20, 2020, WHO officially named this disease caused by the new coronavirus SARS-CoV-2 as COVID-19 [6]. As of March 1, 2020, a total of 80,369 confirmed cases and 2,915 death cases with COVID-19 infection were reported from 31 provinces (autonomous regions, municipalities) in China [2].

To date, in the first forty days since identification of the COVID-19 (Jan 8th^-^Mar 1th, 2020), considerable new knowledge about this coronavirus have been generated, such as person-to-person transmission [7], clinical characteristic [8-10], prediction on the incubation period, basic reproductive number, total case number, and epidemic trend [11, 12]. However, key question about the CFR, an important epidemiological indicator for reflecting and predicting the severity of the disease, of the COVID-19 epidemic remains unsolved. Timely and accurate estimates of the CFR are critical for predicting the outbreak dynamics and health care capacity needed, to tailor appropriate and effective measures for disease control, public safety, and allocation of health resources.

At the early stage of COVID-19 outbreaks, given the limited and incomplete data (for the epidemic is still ongoing), the CFR is a crude estimated as by dividing the number of known deaths by the number of confirmed cases with COVID-19 in previous papers [8-10]. Nevertheless, these results are subject to large errors because future deaths are not taken into account, instead, which might be off by orders of magnitude. Diagnosis of viral infection will develop to two competing risk outcomes, recovery or death, by days to weeks and the denominator of CFR, the number of confirmed cases, at the cross-sectional level should consider the loss of those recovered [13]. Hence in this study we aim to provide a precise estimate of the case fatality ratio of COVID-19 epidemic at present.

## Methods

### Data sources

We extracted the daily released epidemic report published by the National Health Commission P. R. China from 20 Jan, 2020, to 1 March, 2020 [14]. For each day, the number of new admissions, cumulative number of cases, number of new deaths, cumulative number of deaths, newly cured patients, and cumulative number of cured patients were obtained.

### Statistical analysis

Although it did not include any individual patient data, the CFR estimated using the competing risks model in the survival analysis [15]. Following the standard non-parametric competing risks theory, the daily hazards for dying and cure, treated as competing risks, are each estimated, and the other endpoint are treated as censoring.

For each day i, we captured the number of new admissions (ni), cumulative number of cases (Ni), number of new deaths (di), cumulative number of deaths (Di), newly cured patients (ci), and cumulative number of cured patients (Ci).

Other parameters listed below:

t: the survival time during the admission-to-cure or death;

J: 1denoted for death, 2 denoted for cure;

λ: 1 denoted for the daily hazards for dying, 2 denoted for the daily hazards for curing, other endpoints are censored;

mi: equaled to Ni − Di – Ci, denoted for the number of patients in hospital;

ai: equaled to (mi−1 + mi)/2, denoted for the average number of patients in hospital (at risk) for each day I;

λi1, equaled to di/ai, denoted for the daily hazards for death;

λi2, equaled to ci/ai, denoted for the daily hazards for cure;

Λ1(k), equaled to 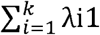, denoted for the cumulative hazards for death up to k days; Λ2(k), equaled to 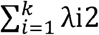, denoted for the cumulative hazards for cure up to k days;

θk, equaled to Λk2/Λk1, denoted for the cure-death hazard ratio for each day k.

P(J), J=1 denoted for the ultimate case fatality ratio, J=2 denoted for the ultimate case cure rate.

For up to day k, the cumulative hazards for death, cure, and cure-death hazard ratio were obtain. If the ratio is constant θ over the study period, then it follows that P(J=2)=θP(J=1). Since P(J=1)+P(J=2)=1, we P(J=1)=1/(1+θ). Therefore, if the ratio is a constant θ during a period then the ultimate CFR in that period is defined as 1/(1+θ). To investigate the relationship between cure-death hazards was linear or non-linear, the cure-death hazard plot was shown. If non-linear relationship was discovered, joinpoint regression will be used [16].

## Results

### Data description

As of March 1, 2020, a total of 80,369 cases were diagnosed as COVID-19 in China. Total number of death cases and cured cases of COVID-19 were 2,803 and 33,757 in Hubei province, and 112 and 10,761 in other provinces in China, respectively. The number of deaths and cures in Hubei, including Wuhan, accounted for 96.2% and 75.8% of the whole country, but the proportion of deaths and cures was quite different, which reminded us to separate the fatality rate of Hubei and other areas (Fig. 1).

**Fig. 1.**
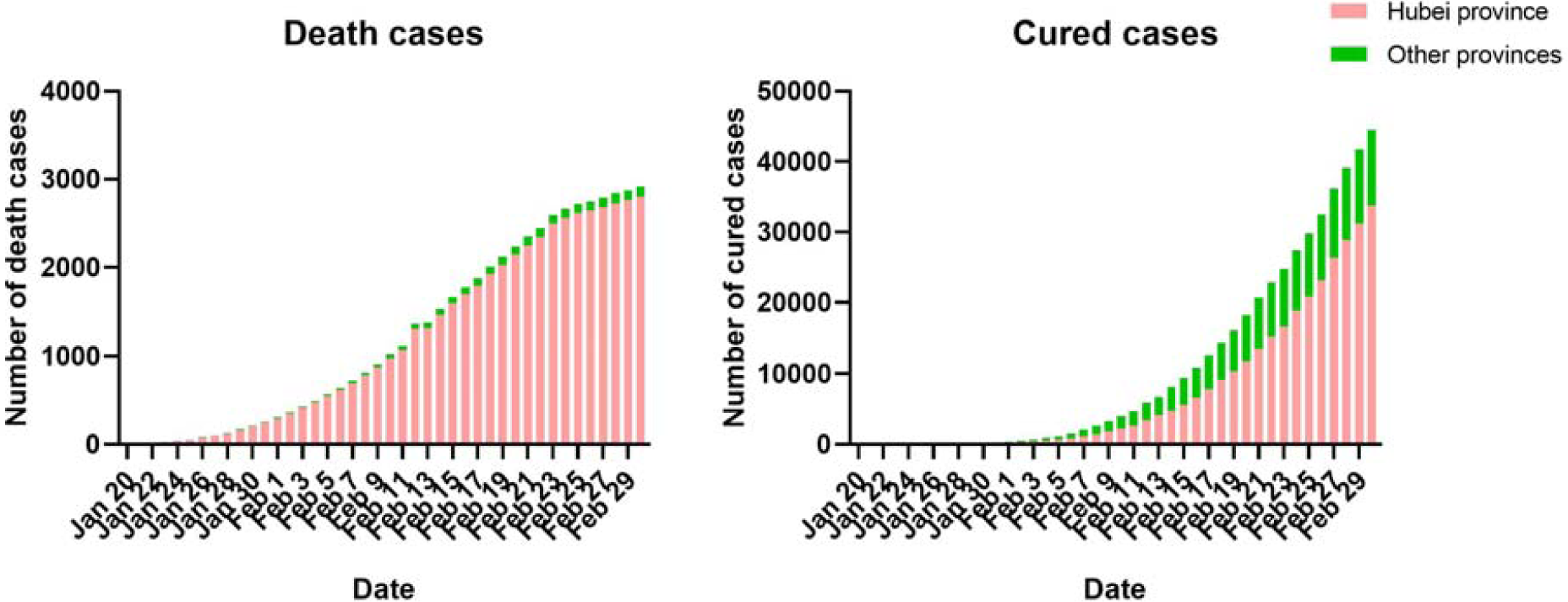
Cumulative numbers of death and cured cases in Hubei and other province.

### CFR estimation of the COVID-19

In the joinpoint regression, the relationship between cure-death hazards were nonlinear for both Hubei and other provinces (Fig. 2). With the slope (cumulative cured hazard/cumulative death hazard) of joinpoint fitting increased, the corresponding CFR estimation gradually decreased. Our CFR estimation was calculated based on the slope of the last piece in joinpoint regression model, which reflected the most recent trend of the epidemic. As shown in Fig. 3, the CFR of COVID-19 were estimated to be 70.9% (95% CI: 66.8%-75.6%) during Jan 20-Feb 2, 20.2% (18.6%-22.1%) during Feb 3-14, 6.9% (6.4%-7.4%) during Feb 15-23, 1.5% (1.4%-1.6%) during Feb 24-March 1 in Hubei province, and 20.3% (17.0%-25.3%) during Jan 20-28, 1.9% (1.8%-2.1%) during Jan 29-Feb 12, 0.9% (0.8%-1.1%) during Feb 13-18, 0.4% (0.4%-0.5%) during Feb 19-March 1 in other provinces of China, respectively.

**Fig. 2.**
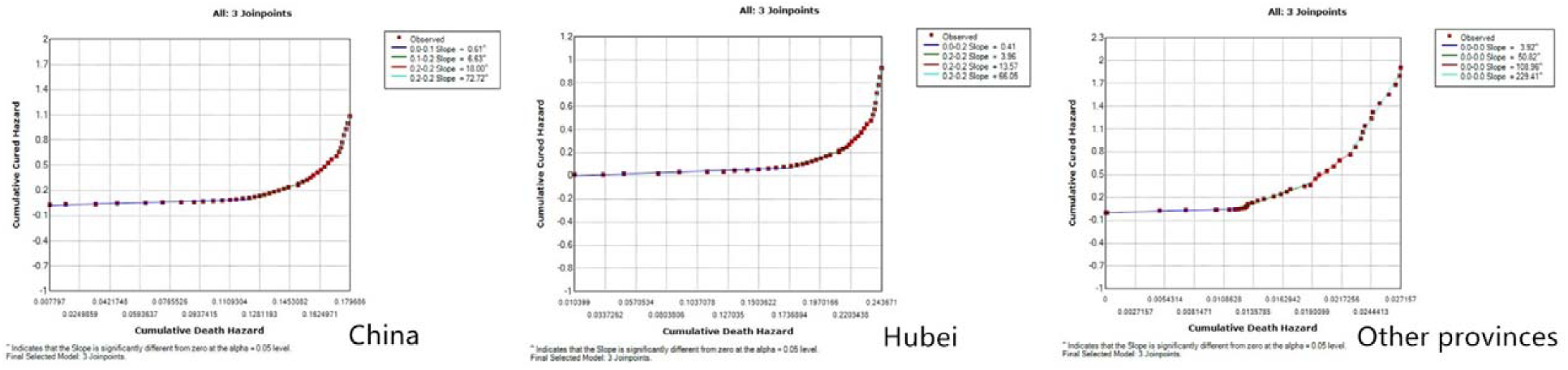
Joinpoint regression of cumulative cured hazard vs. cumulative death hazard.

**Fig. 3.**
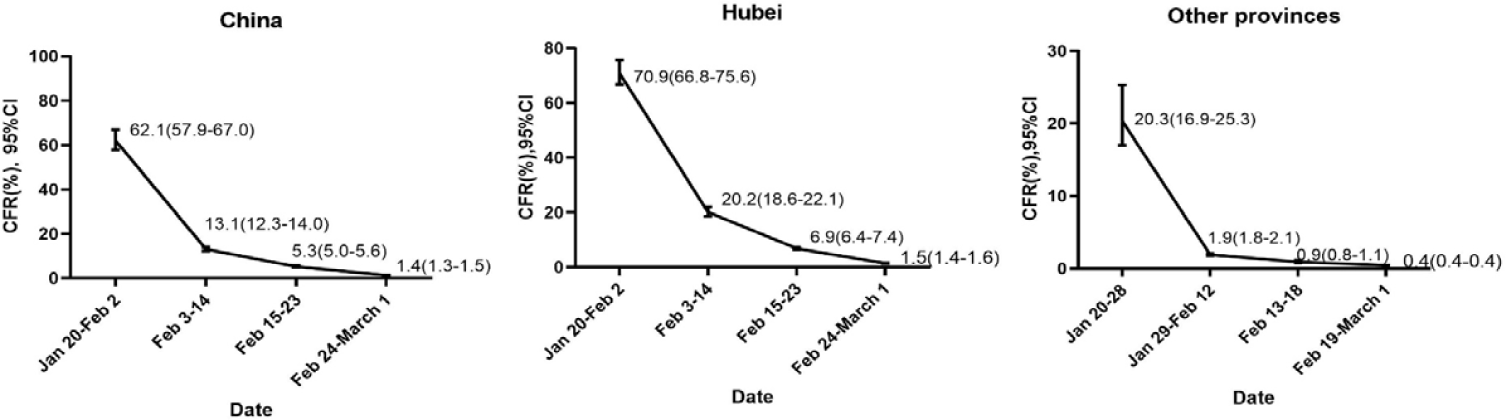
Estimated CFR of COVID-19 during different time periods in China, Hubei and other provinces.

## Discussion

Based on public available daily data, we found that the CFR of COVID-19 changes with time and decreases continuously both in Hubei and other provinces.

The most current estimate of the CFR of 1.5% in Hubei for COVID-19 is lower than that of SARS-CoV (9.2%) and that of MERS-CoV (34.4%) [17]. Consistent with our results, Chinese researchers reported comparable CFR estimations of 11% among 99 patients in Wuhan Jinyintan Hospital [9] and 15% among 41 patients in Wuhan [10]. However, others reported markedly lower CFR (1.4%) among 1,099 patients from 552 hospitals in 31 provinces/provincial municipalities and 4.3% among 138 patients in Wuhan Zhongnan Hospital [8]. It should be noted that these CFR estimates were cross-sectional, which were calculated by dividing the number of known deaths by the number of confirmed cases at certain time point[18]. As contrast, WHO Collaborating Centre for Infectious Disease Modelling and MRC Centre for Global Infectious Disease Analysis published their 4th edition Report: Severity of 2019-novel coronavirus (nCoV), which showed a markedly higher estimation of CFR to be 18% (95% CI: 11%-81%) for cases detected in Hubei [19].

This estimated CFR of COVID-19 was much higher in Hubei Province than that of other area in China. Several possible reasons could be able to explain this discrepancy. Firstly, high proportion of severe cases among the confirmed patients potentially contributes to high proportion of death in Hubei Province [8-10]. Patients with serious symptoms were prioritized to be admitted to the hospital. Secondly, insufficient medical resources including clinicians, nurses as well as facilities, including diagnosis toolkits at the outbreak of COVID-19 in Hubei directly resulted the deterioration of disease course. It is reasonable to assume that huge amount of mild cases or early-stage cases have developed into more serious conditions before they were able to be admitted and appropriately treated. Thirdly, under-detection of mild or asymptomatic cases also resulted in higher CFR, which may be further aggravated after the outbreak, due to that the daily capacity of virus testing in Hubei has already reached the limit. Fourthly, as the COVID-19 initially outbroke in Wuhan, Hubei, other regions of China and foreign countries gained critical time to develop epidemic control strategy and reallocate medical resources. However, this delay of epidemic in other regions and foreign countries may also result in the delay of fatal cases arising and reporting, which rendered the lower CFR in other regions of China comparing with Hubei Province.

We observed obvious decreasing trends of CFR for COVID-19, with distinct turning points for in Hubei and other areas respectively. From Jan 23, 2020, several unprecedentedly strict and effective measures were carried out, especially the lockdown of Wuhan, Hubei, including suspended public transportation, forbidden pubic gathering, postponed school and enterprise opening, and furnished kinds of medical resources to prevent further disease transmission [20]. Based on our estimation and recently released data, we concluded that the CFR would follow a downwards trend.

Actually, the extent of underreporting varied over time, cities and countries [13]. In fact, in order to break the capacity limit of virus lab testing and achieve a better control of this epidemic, Hubei Province has included the clinically diagnosed cases into the confirmed cases to be published starting from February 12, 2020. Clinically diagnosed cases are confirmed by a combination of travel history, symptoms and CT image rather than the virus lab test. So there are 14,840 new confirmed cases (including 13,332 clinically diagnosed cases) in Hubei province was reported in February 12, 2020, abided by the diagnosis and treatment scheme for the new coronavirus infections (pilot version 5) [21]. In this case, our CFR estimated will decrease due to the update of diagnosis standard.

The competing risk model, for estimating the CFR in the early stage of COVID-19 epidemic, is applicable to any disease for which the final outcome is not known for a proportion of patients [18]. Using published daily summary data when individual data is not available, provides a powerful sample to estimate CFR and to predict the future trend of disease epidemic.

Nevertheless, several limitations should be considered. First, our analyses were based on public summary data with a lack of individual level of time to death or cure, characteristics at baseline, such as age, gender and chronic disease status. Therefore, the heterogeneity of CFR among the subgroups is not able to be investigated. Neither is it easy to find the most susceptible population of COVID-19 to whom better protections should be provided. However, previous studies have found that males older than 65 years with multiple comorbidities such as cardiovascular diseases are the most vulnerable population than others, suffering with both the highest incidence of confirmed patients and the highest CFR [8-10]. Second, currently in our study, it is difficult to adjust the influence of the lag of the real case numbers resulting from insufficient medical resources in Hubei Province. With the development of the epidemic, this lag will be alleviated and then we can get a more precise idea of the severity of this COVID-19 epidemic.

## Conclusion

Based on analyses of public data, we found that the adjusted CFR in Hubei was much higher than that of other regions in China, over 3 times in all estimation. The CFR would follow a downwards trend based on our estimation and recently released data. Nevertheless, at early stage of outbreak, CFR estimates should be viewed cautiously because of limited data source on true onset and recovery time.

## Data Availability

All data, models, and code generated or used during the study appear in the submitted article.

## Notes

### Competing Interest Statement

The authors have declared no competing interest.

### Funding Statement

No funding was received for the research reported in the article.

